# Divergent social patterning of directly measured environmental exposures across Rhode Island communities

**DOI:** 10.64898/2026.08.20.26360933

**Authors:** Erica D. Walker, Sai Venkat Mandalapu, Sage Lefebvre

## Abstract

**Background:** Environmental noise and air pollution are both shaped by road traffic and the built environment, and exposure assessment increasingly folds them into composite indices or proxies both by traffic exposure. Whether the two share a social distribution has rarely been tested against direct measurement of several exposures in the same communities, and community noise is almost always characterized by A-weighted levels alone, which discount low-frequency energy.

**Methods:** At 176 sites across Rhode Island, spanning the contiguous urban area of Providence, Central Falls, and Pawtucket together with four rural municipalities, we measured the acoustic environment under A- and C-weighting (LAeq, LCeq), fine particulate matter (PM2.5), night- time illuminance, and relative humidity across four session types over roughly one year (704 site-sessions). Exposures were linked to census-tract composition (American Community Survey), and mixed-effects models were fitted for each of eight area-level markers of disadvantage, adjusting for campaign and session. Relative humidity was carried through the identical model as a negative control.

**Results:** A-weighted noise was consistently higher in more disadvantaged tracts, rising with non-White, poverty, renter, and no-vehicle shares and falling with income and older-resident share (six of eight markers significant; 1.3 to 1.8 dBA per standard deviation; 6.6 dBA between the least and most racially diverse neighborhoods). C-weighted levels followed the same gradient on every marker and exceeded their A-weighted counterparts at block-group scale for renter occupancy and vehicle absence. Night-time illuminance was also socially patterned, whereas short-term PM2.5 was roughly an order of magnitude weaker and relative humidity showed no gradient. The acoustic gradient persisted within the urban core alone.

**Conclusions:** Measured burden was carried by the acoustic environment, including its low-frequency component, and by night-time light, not by short-term particulates. The exposure metric and the averaging time determine which disparities are visible at all.

**Impact statement:** Composite environmental indices and traffic surrogates assume that a neighborhood burdened on one exposure is burdened on all. Measuring noise, particulate matter, and night-time light directly at the same 176 sites shows otherwise: the social gradient was carried by the acoustic environment and by night-time light, not by short-term particulates. Reporting C-weighted alongside A-weighted levels, which to our knowledge has not previously been examined as a distributional measure, shows that the low-frequency burden falls on the same disadvantaged communities and is understated by the conventional metric. Exposure assessment choices therefore determine which inequalities become visible.

## 1. Introduction

Geographic location is a primary determinant of individual environmental exposure. Decades of work on the unequal distribution of environmental hazards have shown that lower-income communities and communities of color often carry a heavier burden of pollution, industrial land use, and other environmental stressors than wealthier and whiter neighborhoods [1,2,3]. Two exposures that figure prominently in this literature are air pollution and environmental noise, and both have well-documented consequences for health. Long-term exposure to fine particulate matter (PM2.5) is among the most firmly established environmental risk factors for cardiovascular and respiratory disease and premature death [3]. Environmental noise, long treated as a nuisance rather than a hazard, is now recognized by the World Health Organization as a cause of sleep disturbance, cardiovascular disease, and other harms [4,5,6,7,8].

Road traffic is a major source of both noise and combustion-related air pollutants, inviting the assumption that the two exposures represent different manifestations of the same underlying environmental disadvantage. Under this view, the same neighborhoods are expected to be simultaneously loud, polluted, and otherwise environmentally burdened. This expectation is reflected in the increasing use of composite environmental indices and in the common practice of using traffic proximity as a surrogate for multiple environmental exposures. Whether the assumption holds has important practical consequences. If noise and air pollution are distributed along different social and spatial gradients, collapsing them into a single environmental burden could misdirect environmental-justice monitoring toward the wrong exposure and misclassify exposure in epidemiological studies that use one as a proxy for the other. Yet co-located measurement studies have consistently found that traffic noise and traffic-related air pollution are only moderately correlated, and that roadway proximity is an imperfect surrogate for both [9,10]. Rather than resolving this issue, the exposome framework, which seeks to characterize the totality of environmental exposures, heightens the need to determine whether ostensibly related exposures actually share the same social and spatial patterning; if they do not, treating them as interchangeable obscures potentially important differences in environmental risk [11].

There are good physical reasons to expect noise and PM2.5 to be distributed differently. Community noise is dominated by road traffic, structured by land use, and stable enough at a given location to be predicted from fixed features of the built environment [12,13]. Land-use regression models built from physical predictors such as road type, traffic volume, building density, and vegetation routinely explain most of the spatial variation in measured noise across cities in North America and Europe [12,13,14,15]. In this sense, noise is a property of place: it is built into the configuration of roads, buildings, and industry around a site and tends to persist. Fine particulate matter behaves differently within an urban area. A large share of urban PM2.5 is regional and secondary rather than emitted at the curb, so concentrations tend toward spatial homogeneity across a metropolitan area while shifting from day to day with the weather [16,17]. Short-term measured PM2.5, in particular, reflects the meteorology of the measurement window as much as the characteristics of the location.

These contrasting spatial logics have a direct consequence for the study of environmental inequality. If noise is fixed in place while short-term PM2.5 floats with the weather, then the two need not be socially patterned in the same way, even where both ultimately derive from traffic. A substantial body of work has documented that noise exposure is socially patterned. In the contiguous United States, modeled noise is higher in census block groups with larger non-White and lower-income populations, and the disparity is amplified by residential segregation [18]. Contemporary transportation-noise disparities track historical redlining [19], and tract-level analyses find sharply elevated odds of high noise exposure among racial and ethnic minority residents [20]. Reviews of European and North American studies report similar, if heterogeneous, gradients [21,22], and a recent national study in the Netherlands found environmental noise concentrated in socioeconomically less privileged neighborhoods [23]. A recurring limitation of this literature, however, is reliance on modeled rather than measured noise, and the treatment of noise in isolation from air pollution. Few studies have measured both exposures directly, in the same communities, and asked whether they follow the same social gradient.

A further limitation is that this literature characterizes noise almost entirely through A-weighted levels. A-weighting approximates the frequency sensitivity of human hearing and therefore discounts low-frequency energy, with the result that A-weighted levels underestimate the response to noise that is rich in low frequencies [24]. The difference between C-weighted and A-weighted levels of the same sound indexes how much low-frequency energy that sound contains; low-frequency noise is both less readily attenuated by building facades and less amenable to conventional barrier mitigation than higher-frequency sound [24]. These properties matter for the distributional question. If the low-frequency component of the acoustic environment is concentrated in the same communities that carry the highest overall levels, then those communities face a burden that is systematically understated by the metric conventionally used to describe it, and that penetrates the housing in which they live. C-weighting is rarely reported in community noise assessment and, to our knowledge, has not previously been examined as a distributional measure. We therefore report C-weighted alongside A-weighted levels throughout.

A comparison confined to noise and particulate matter would also leave open whether any gradient it revealed was specific to those exposures or common to anything measurable in a disadvantaged neighborhood. Two further environmental channels allow that question to be settled directly. Night-time outdoor light is a plausible exposure in its own right: artificial light at night disrupts circadian rhythm and sleep, and a large United States cohort study found the association between outdoor light at night and short sleep duration to be stronger in higher-poverty neighborhoods [25]. Relative humidity, by contrast, has no plausible mechanism linking it to neighborhood composition and therefore serves as a negative control: were a social gradient to appear in every quantity measured, including that one, the pattern would point to a methodological artifact rather than to a structured difference in exposure.

We address this gap by directly comparing the social patterning of measured environmental exposures across Rhode Island communities. Using standardized protocols, the acoustic environment under A- and C-weighting, fine particulate matter, night-time illuminance, and relative humidity were measured at 176 sites spanning the contiguous urban area of Providence, Central Falls, and Pawtucket together with four rural municipalities, with each site visited across four session types defined by day of week and time of day over roughly one year. We linked site-level exposures to census-tract socioeconomic and racial composition to test whether these exposures exhibit similar social gradients or instead identify different communities as environmentally burdened. This is at bottom an exposure-assessment question, and it is consequential for three audiences: environmental epidemiologists, for whom one exposure standing in for another is a source of misclassification; those who build and use composite environmental indices and screening tools, whose output determines which communities are identified as overburdened; and state and municipal agencies responsible for noise and land-use decisions, for whom the addressable target depends on which exposure carries the disparity. We interpret our findings within an environmental-justice framework, with specific concerns for the distribution of measured environmental exposure across communities that differ in socioeconomic and racial composition. Throughout, we interpret these associations ecologically, describing the characteristics of places rather than of individuals.

## 2. Methods

### 2.1. Study area and design

Measurements were collected at 176 sites across Rhode Island, United States. The sites were drawn from two field campaigns. The first covered the contiguous urban area comprising Providence, Central Falls, and Pawtucket, contributing 144 sites; the second campaign focused on four rural municipalities and contributed 32 sites.

Rhode Island and these particular places were selected deliberately rather than for convenience. The state is small enough that a single coordinated field effort can span its full settlement gradient, from densely developed urban neighborhoods to rural communities, yet it contains pronounced socioeconomic heterogeneity across short distances. These three contiguous cities are among the state’s most densely populated and lowest-income municipalities, which also experience substantially higher concentrations of childhood poverty and educational disadvantage than the rest of Rhode Island. The four rural municipalities were selected because they meet Rhode Island’s definition of rural. Because the state’s high population density and small land area make federal rural classifications a poor fit, Rhode Island applies its own rural definition [26]. Sampling both ends of this gradient was necessary to obtain the range of neighborhood composition required to detect social patterning at all.

These two community groups differ in more than present-day socioeconomic composition. The urban core developed as a manufacturing center during industrialization and was settled through successive waves of immigration, leaving a dense, mixed-use fabric of mills, arterial roads, and rail corridors in close proximity to housing; mid-twentieth-century highway construction subsequently routed limited-access roadways through several of these neighborhoods, and parts of the core have more recently experienced reinvestment and demographic change. The rural municipalities developed later and more sparsely, with lower road densities and greater separation between housing and heavy traffic. These histories are not represented in our models, but they are why the two settings differ systematically in the fixed physical features that generate noise. Rhode Island also carries a high chronic respiratory disease burden: the state recorded the highest adult current asthma prevalence in the United States in recent national surveillance, at 12.6% against a state median of 9.8% [27], making the distribution of measured air quality locally salient.

Each site was scheduled for measurement during four session types defined by day of week and time of day: weekday daytime, weekday nighttime, weekend daytime, and weekend nighttime. Daytime sessions covered approximately 07:00 to 19:00 and nighttime sessions approximately 19:00 to 07:00. The campaigns were carried out over roughly one year. Measurements were collected by trained students following a standardized protocol for instrument placement, timing, and data recording.

### 2.2. Exposure measurements

At each site-session, the acoustic environment was characterized by the equivalent continuous sound level recorded over a fixed five-minute sampling interval under three frequency weightings: A-weighted (LAeq, in dBA), C-weighted (LCeq, in dB), and unweighted or Z- weighted (LZeq, in dB). A-weighting is the convention in community noise assessment and approximates the frequency response of human hearing; C-weighting applies far less attenuation below 500 Hz and therefore retains low-frequency energy that A-weighting discards, so that the difference between the two indexes the low-frequency content of the measured sound [24]. We report A- and C-weighted levels together throughout, and treat C-weighting as a distributional measure in its own right rather than as a check on the A-weighted result. Z-weighted levels are reported in the Supplementary Material for completeness (Supplementary Table S7). Air quality was measured with portable sensors reporting fine particulate matter (PM2.5) together with coarse particulate matter (PM10), the air quality index, and gaseous pollutants (total volatile organic compounds and formaldehyde). Night-time outdoor illuminance was recorded in footcandles at ten-second intervals during night sessions and summarized as the session mean; because it is defined only for night sessions, the illuminance models rest on the night-time subset of site-sessions. This measure captures total outdoor illuminance after dark and therefore includes any natural contribution from moonlight and starlight alongside artificial sources; it is a measure of the night-time light environment rather than of artificial light at night specifically. Relative humidity was recorded by the same air-quality sensor and is included as a negative control. Site coordinates were recorded in the field and checked before analysis; coordinates with sign or formatting errors were corrected against the recorded location. For each site we summarized exposure as the mean across that site’s sessions, giving one acoustic value and one PM2.5 value per site for descriptive purposes, while session-level observations were retained for the mixed-model analysis described below. PM2.5 was log-transformed for analysis to reduce right-skew. The portable sensors used here are suited to relative comparison across sites and sessions rather than to regulatory-grade absolute concentration, and all analyses rely on relative differences across locations and times.

### 2.3. Neighborhood socioeconomic and racial composition

Each site was assigned to a census tract by geocoding its coordinates against the United States Census Bureau geographic reference files. We re-derived all tract and block-group assignments directly from coordinates rather than relying on previously stored codes, and we verified the derived tract assignments against independently recorded values. For each tract we obtained socioeconomic and racial composition from the American Community Survey (ACS) 2023 five-year estimates, computed from the published count tables with appropriate denominators. We used eight area-level markers of neighborhood composition: the percentage of residents who were non-White (defined as one minus the non-Hispanic White share), the percentage below 200% of the federal poverty level, the percentage with less than a high-school education among adults aged 25 and over, the percentage of housing units that were renter-occupied, the percentage of households with no vehicle available, the percentage of residents under 18, the percentage of residents 65 and over, and median household income.

These eight markers were chosen to span the dimensions of neighborhood composition most often used in distributional analyses of environmental exposure, and they overlap substantially with the criteria Rhode Island applies when designating an Environmental Justice Focus Area [28], which a census tract meets if any of several thresholds is satisfied: a median household income no greater than 65% of the statewide median, a minority population of at least 40%, at least 25% of households lacking English-language proficiency, or a combination of a minority population of at least 25% with a municipal median income no greater than 150% of the statewide figure. Our income and racial-composition markers correspond closely to the first two criteria. The one criterion we could not represent is English-language proficiency, which we did not include because the corresponding ACS estimates at tract level carry margins of error too large to support the site-level linkage used here; we note this as a gap rather than a judgment about its relevance.

Census tracts were the primary unit of analysis. We treated tract-level estimates as the more stable choice because ACS estimates at finer geographies carry larger margins of error, and at the census-tract scale these margins are already on average considerably larger than those of the long-form census they replaced [29,30]. For this reason we relied chiefly on percentage-based composition measures rather than on median income, whose margins of error are larger, and we report income associations as descriptive. Block-group estimates were retained for a sensitivity analysis. The use of area-level composition to describe the social environment of a site is a standard but consequential choice; associations estimated at the area level describe places, not the individuals within them, and are subject to the ecological fallacy and to the modifiable areal unit problem, whereby results can depend on the size and shape of the units chosen [31,32]. We return to these limitations in the Discussion.

### 2.4. Statistical analysis

We modeled each exposure as a function of neighborhood composition using linear mixed-effects models. Five measured outcomes were carried through the identical model structure: A-weighted noise, C-weighted noise, log-transformed PM2.5, night-time illuminance, and relative humidity, with Z-weighted noise reported in the Supplementary Material. Applying one specification to every measured channel, rather than to the exposures of primary interest alone, means that null results are reported on the same footing as positive ones and that the specificity of any gradient can be assessed directly. For each of the eight composition markers and each outcome, we fitted a separate model with the standardized composition marker as the predictor, a random intercept for site to account for repeated visits, a fixed effect for session type, and a fixed effect for campaign to account for the pooling of the two field efforts. Fitting one marker at a time, rather than entering all eight together, avoids the strong collinearity among neighborhood composition measures and yields associations that are directly interpretable; it does not attempt to separate the unique contribution of each marker, which the data do not support. Predictors were standardized to zero mean and unit variance, so each coefficient expresses the change in exposure per standard deviation of the composition marker, in decibels for noise and in log units for PM2.5. This pooled, campaign-adjusted, tract-level model is our primary specification.

We assessed the robustness of the findings in three ways. First, to confirm that the results were not an artifact of the repeated-measures structure, we collapsed each exposure to a single site-level mean and refitted the eight models with ordinary least squares. Second, to test the dependence on spatial scale, we refitted the primary models using block-group rather than tract composition. Third, to test whether the noise pattern was merely an urban-versus-suburban contrast introduced by pooling two dissimilar campaigns, we refitted the models within the Providence urban core alone, where neighborhood composition still varies substantially but the urban-suburban distinction does not apply. All analyses were conducted in R. We interpret the consistency of direction and magnitude across these specifications, rather than any single p-value, as the basis for our conclusions.

## 3. Results

### 3.1. Exposure and neighborhood composition across the sample

The 176 sites spanned a wide range of neighborhood composition (Table 1). The percentage of non-White residents ranged from 3.8% to 97.4% across the sampled tracts (median 40.2%, interquartile range 17.6% to 72.1%), and the other markers were similarly broad: the percentage below 200% of poverty ranged from 3.0% to 71.1%, the percentage of renters from 4.2% to 94.8%, and median household income from about $16,000 to $229,000. Measured noise also varied widely, with a site-level median LAeq of 57.5 dBA (interquartile range 51.9 to 63.1, full range 40.9 to 75.7) and a median LCeq of 70.2 dB (65.1 to 73.7, full range 51.5 to 97.5). The C-weighted median therefore exceeded the A-weighted median by 12.7 dB across these sites, indicating that a substantial share of the acoustic energy present lay in the low frequencies that A-weighting discounts. Measured PM2.5 was low and spanned a narrow range, with a site-level median of 4.0 µg/m³ (interquartile range 2.6 to 5.0, full range 1.0 to 10.8), consistent with the generally clean regional air during the measurement period. Pooling the urban core (144 sites) and the rural municipalities (32 sites) was central to the design: it produced the range of neighborhood composition needed to detect social gradients, while the within-core analysis described below guards against attributing those gradients to the urban-suburban contrast alone.

**Table 1.** Study design and descriptive statistics across the 176 sites. Exposure values are site-level summaries; neighborhood markers are census-tract values assigned to each site. Values are median (interquartile range) and full range.

| Variable | Median (IQR) | Range |
| --- | --- | --- |
| Sites | 176 | — |
| Providence urban core / rural municipalities | 144 / 32 | — |
| Noise, LAeq (dBA) | 57.5 (51.9–63.1) | 40.9–75.7 |
| Noise, LCeq (dB) | 70.2 (65.1–73.7) | 51.5–97.5 |
| Noise, LZeq (dB) | 73.4 (70.4–77.7) | 57.8–98.4 |
| PM2.5 (µg/m<sup>3</sup>) | 4.0 (2.6–5.0) | 1.0–10.8 |
| % non-White | 40.2 (17.6–72.1) | 3.8–97.4 |
| % below 200% poverty | 30.9 (20.1–49.0) | 3.0–71.1 |
| % renter-occupied | 59.9 (34.1–74.2) | 4.2–94.8 |
| % no vehicle | 16.0 (6.5–22.2) | 1.1–46.6 |
| % less than high school | 10.1 (5.6–19.0) | 0.0–44.3 |
| Median household income (USD) | 72,779 (54,747–98,125) | 16,448–229,107 |
| % over 65 | 12.8 (10.5–21.6) | 3.3–45.2 |
| % under 18 | 17.8 (6.8–22.0) | 0.5–40.7 |

### 3.2. Neighborhood disadvantage tracks measured noise

In the primary specification, measured noise was consistently and significantly higher in more disadvantaged tracts (Table 2, Fig. 1; full coefficients and standard errors in Supplementary Table S1). Per standard deviation of neighborhood composition, site noise rose by 1.41 dBA with the percentage of non-White residents (p = 0.017), 1.26 dBA with the percentage below 200% of poverty (p = 0.028), 1.77 dBA with the percentage of renters (p = 0.004), and 1.75 dBA with the percentage of households without a vehicle (p = 0.001). Noise fell where advantage was greater, declining by 1.34 dBA per standard deviation of median income (p = 0.013) and by 1.54 dBA per standard deviation of the percentage of residents aged 65 and over (p = 0.006). The two remaining markers pointed in the expected directions but did not reach significance: the percentage with less than a high-school education (0.50 dBA, p = 0.34) and the percentage of residents under 18 (−0.81 dBA, p = 0.10). Six of the eight markers were thus significant, and all eight were oriented consistently, with greater disadvantage associated with higher A-weighted noise. The corresponding C-weighted associations are reported in Section 3.3.

**Figure 1.**
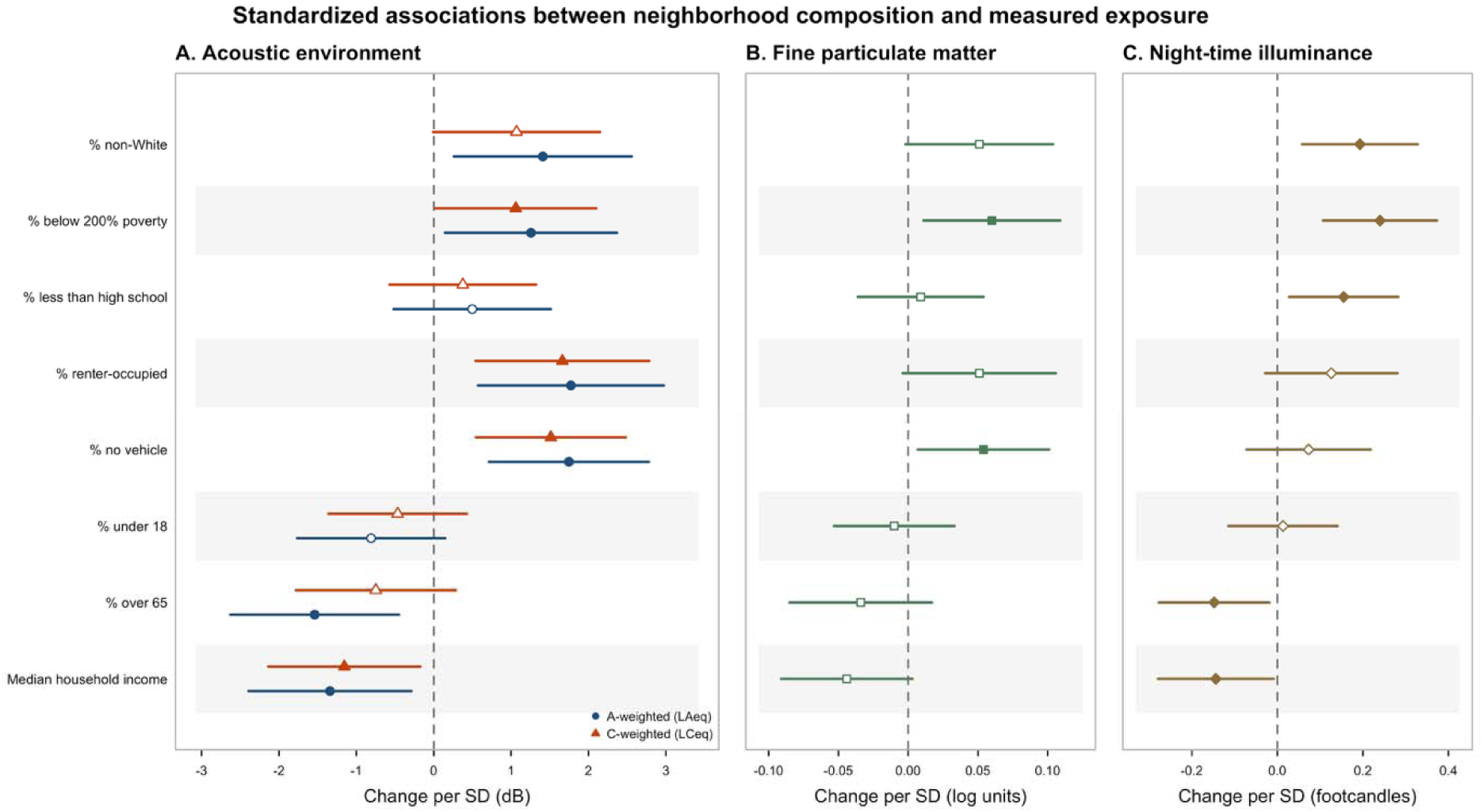
Standardized associations between neighborhood composition and each measured exposure for eight area-level markers, estimated from campaign- and session-adjusted mixed models across 176 sites (704 site-sessions). Points are coefficients per standard deviation of the composition marker and horizontal lines are 95% confidence intervals; filled markers indicate p < 0.05. Panel A shows the acoustic environment under both frequency weightings on a common decibel axis, A-weighted (LAeq) and C-weighted (LCeq); Panel B shows log-transformed PM2.5; Panel C shows night-time outdoor illuminance. The acoustic associations are substantial and, for most markers, distinguishable from zero, with greater disadvantage associated with higher levels under both weightings, and the C-weighted associations track the A-weighted associations in direction on every marker. Illuminance associations are positive and distinguishable from zero for five of the eight markers, whereas PM2.5 associations cluster near zero. Relative humidity, included as a negative control, is reported in Supplementary Table S8.

**Table 2.** Standardized associations between neighborhood composition and each measured exposure, primary specification (pooled, tract-level, campaign- and session-adjusted mixed models). Coefficients are per standard deviation of the composition marker: dBA for A-weighted noise, dB for C-weighted noise, log units for PM2.5, footcandles for night-time illuminance, and percentage points for relative humidity. Standard errors for the A-weighted noise and PM2.5 models are given in Supplementary Table S1; C-weighted, Z-weighted, illuminance, and humidity coefficients across all specifications are given in Supplementary Tables S6 to S8.

| Neighborhood marker | LAeq $\beta$ (p) | LCeq $\beta$ (p) | log PM2.5 $\beta$ (p) | Illuminance $\beta$ (p) | Humidity $\beta$ (p) |
| --- | --- | --- | --- | --- | --- |
| % non-White | 1.409 (0.017) | 1.069 (0.053) | 0.051 (0.057) | 0.193 (0.006) | 1.076 (0.094) |
| % below 200% poverty | 1.256 (0.028) | 1.060 (0.047) | 0.060 (0.020) | 0.240 (<0.001) | 0.662 (0.281) |
| % less than high school | 0.497 (0.339) | 0.375 (0.439) | 0.009 (0.716) | 0.155 (0.018) | 0.500 (0.372) |
| % renter-occupied | 1.772 (0.004) | 1.662 (0.004) | 0.051 (0.071) | 0.126 (0.111) | 0.199 (0.766) |
| % no vehicle | 1.746 (0.001) | 1.513 (0.003) | 0.054 (0.025) | 0.073 (0.325) | -0.077 (0.894) |
| % under 18 | -0.810 (0.099) | -0.467 (0.309) | -0.010 (0.666) | 0.013 (0.844) | 1.167 (0.027) |
| % over 65 | -1.540 (0.006) | -0.750 (0.156) | -0.034 (0.196) | -0.148 (0.027) | 0.156 (0.803) |
| Median household income | -1.342 (0.013) | -1.156 (0.022) | -0.044 (0.069) | -0.144 (0.038) | -0.424 (0.463) |

These standardized coefficients correspond to substantial differences across the range of neighborhoods we sampled. Because each marker’s spread is wide, the shift from a relatively advantaged to a relatively disadvantaged tract spans roughly two interquartile ranges, and the noise contrast across a single interquartile range is already on the order of 2 to 3 dBA: about 2.7 dBA across the interquartile range of the percentage of non-White residents, 2.9 dBA across that of renters, 2.2 dBA across that of poverty, and 2.1 dBA across that of vehicle absence.

Comparing the extremes makes the gradient more concrete still: sites in the tracts in the lowest decile of non-White population averaged 51.7 dBA, while those in the highest decile averaged 58.3 dBA, a difference of 6.6 dBA between the least and most racially diverse neighborhoods in the sample. The site-level association with each marker is shown in Fig. 2, and the geographic distribution of measured noise relative to tract racial composition in Fig. 3.

**Figure 2.**
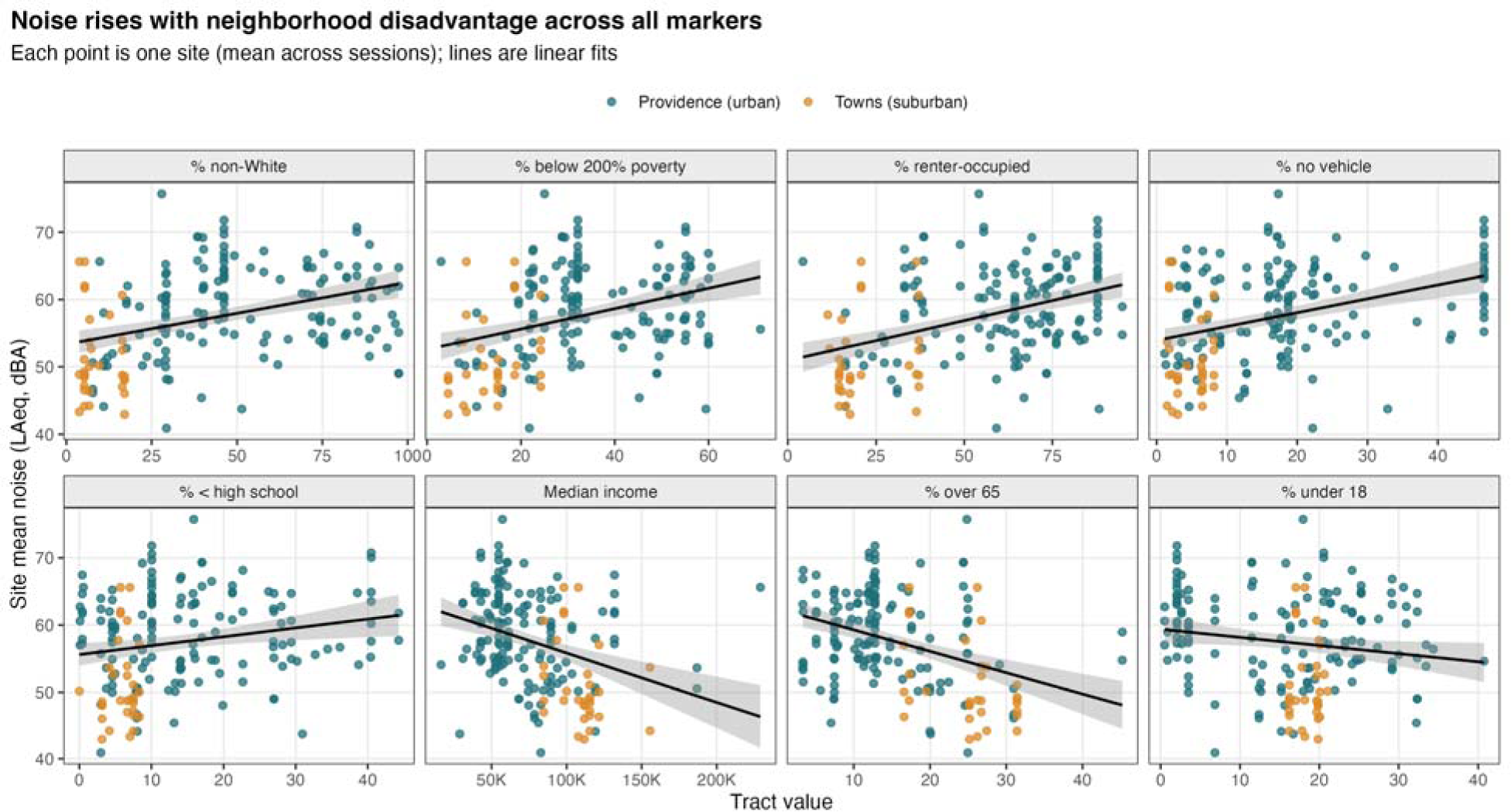
Site-level noise (LAeq) plotted against each of the eight neighborhood composition markers, with points colored by campaign (Providence urban core; rural municipalities) and linear fits overlaid. Noise rises with the percentage of non-White residents, poverty, renter occupancy, and vehicle absence, and falls with income and the percentage of older residents.

**Figure 3.**
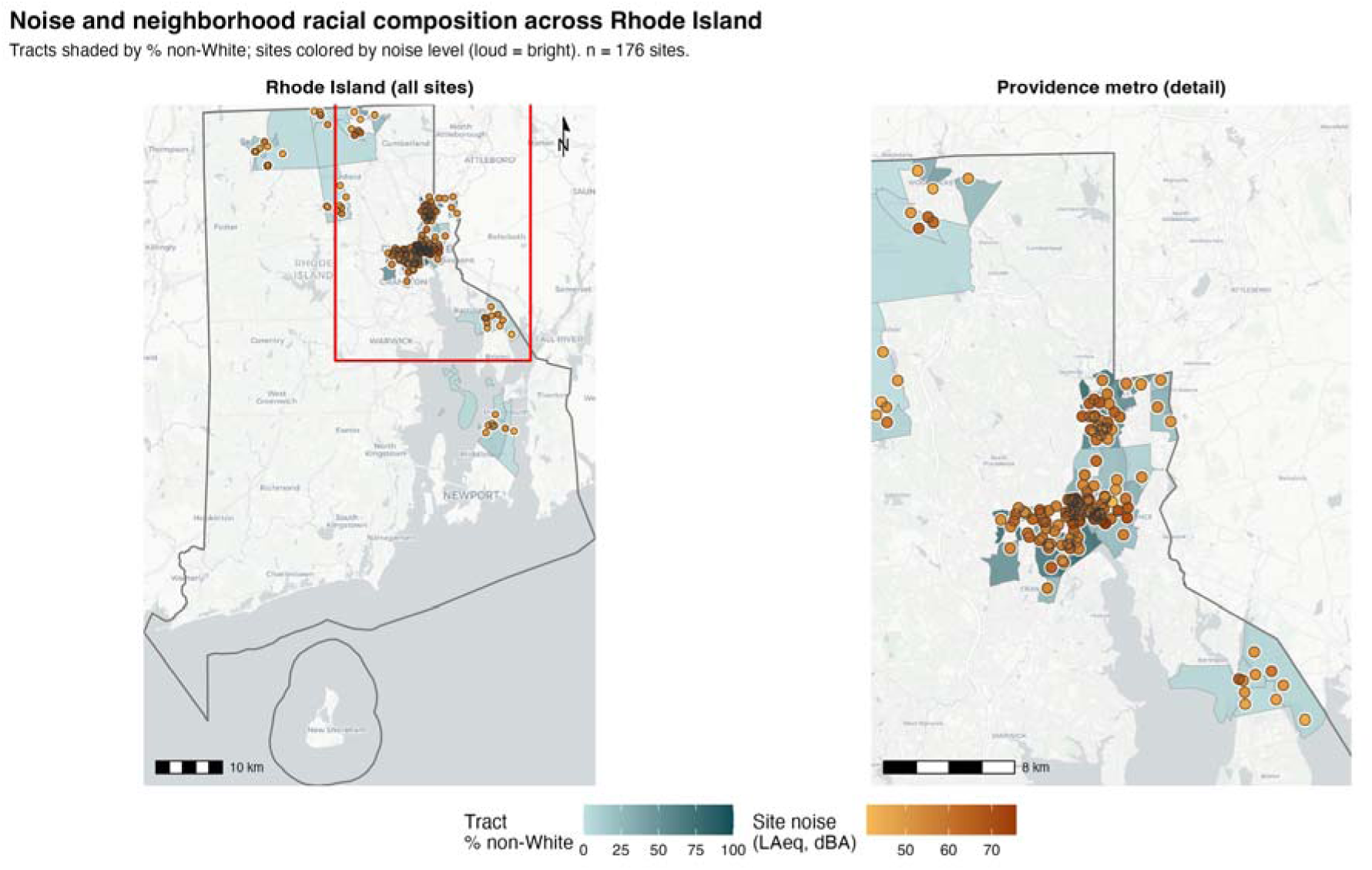
Measured noise and neighborhood racial composition across Rhode Island. Census tracts are shaded by the percentage of non-White residents and monitoring sites are colored by measured noise level, with the full state shown at left and the Providence urban core, where sites are concentrated, shown at right. Louder sites tend to fall in tracts with higher non-White population shares.

### 3.3. The low-frequency burden follows the same social gradient

C-weighted levels, which retain the low-frequency energy that A-weighting discounts, were socially patterned in the same direction as A-weighted levels on all eight markers (Table 2, Fig. 1). Per standard deviation of neighborhood composition, LCeq rose by 1.66 dB with the percentage of renters (p = 0.004), 1.51 dB with the percentage of households without a vehicle (p = 0.003), and 1.06 dB with the percentage below 200% of poverty (p = 0.047), and fell by 1.16 dB per standard deviation of median income (p = 0.022). The association with the percentage of non-White residents was of similar magnitude to its A-weighted counterpart but fell just short of conventional significance (1.07 dB, p = 0.053). Four of the eight markers were significant, with a fifth at the threshold (Supplementary Table S6). Translated across the interquartile range of each marker, these correspond to differences of 2.7 dB for renter occupancy, 2.0 dB for the percentage of non-White residents, 1.9 dB for poverty, and 1.8 dB for vehicle absence. Comparing extremes, sites in the least racially diverse decile of tracts averaged 64.4 dB LCeq and those in the most diverse decile 69.8 dB, a difference of 5.5 dB.

The C-weighted gradient was thus present, consistent in direction, and of comparable magnitude to the A-weighted gradient, but somewhat attenuated: it was strongest for the housing- and transport-related markers, renter occupancy and vehicle absence, and weaker than its A-weighted counterpart for racial composition and for the age markers, which lost significance. Two features of the C-weighted result are worth noting because they run counter to the expectation that C-weighting merely reproduces the A-weighted signal with more noise. First, at the block-group scale, where A-weighted associations attenuated, the C-weighted associations with renter occupancy (2.35 dB per standard deviation, p < 0.001) and vehicle absence (1.94 dB, p < 0.001) were larger than their A-weighted counterparts (2.20 dB and 1.72 dB), and the strongest associations in the entire analysis were C-weighted. Second, the C-weighted associations with renter occupancy and vehicle absence were significant in every specification we fitted. The low-frequency component of the acoustic environment was therefore not incidental to the gradient; on the markers that most directly describe housing tenure and transport dependence, it carried it as strongly as the conventional metric, and at finer spatial scale more strongly.

### 3.4. Short-term PM2.5 shows the same directions but far weaker

Short-term measured PM2.5 was associated with neighborhood composition in the same directions as noise but at a much smaller magnitude (Table 2, Fig. 1). Standardized associations for PM2.5 were on the order of 0.05 log units per standard deviation, roughly an order of magnitude smaller than the corresponding noise associations once the difference in units is set aside, and only two of the eight markers reached significance: the percentage below 200% of poverty (0.060 log units, p = 0.020) and the percentage of households without a vehicle (0.054 log units, p = 0.025). The associations with the percentage of non-White residents (p = 0.057), the percentage of renters (p = 0.071), and median income (p = 0.069) approached but did not reach conventional significance, and the remaining markers were clearly non-significant. The directions of the PM2.5 associations echo the noise pattern, which is consistent with both exposures sharing traffic as a partial source, but their weakness indicates that in this setting the social patterning of exposure was carried mainly by noise rather than by short-term particulate concentrations.

### 3.5. Night-time light is socially patterned; humidity is not

Applying the same specification to the remaining measured channels separated them sharply (Table 2). Night-time outdoor illuminance was socially patterned, and in the same direction as noise: it rose by 0.24 footcandles per standard deviation of the percentage below 200% of poverty (p < 0.001), 0.19 with the percentage of non-White residents (p = 0.006), and 0.16 with the percentage of adults with less than a high-school education (p = 0.018), and fell by 0.15 per standard deviation of the percentage of residents aged 65 and over (p = 0.027) and by 0.14 per standard deviation of median income (p = 0.038). Five of the eight markers were significant, and six of eight pointed toward greater illuminance in more disadvantaged tracts (Supplementary Table S8). The three markers that did not reach significance were renter occupancy, vehicle absence, and the percentage under 18. Within the Providence urban core alone the pattern held for poverty (0.21, p = 0.002), racial composition (0.18, p = 0.005), and education (0.16, p = 0.022), so it was not an artifact of pooling the urban core with the rural municipalities.

Relative humidity, included as a negative control, showed no such structure: only one of the eight markers reached significance (the percentage under 18, p = 0.027), which is consistent with chance across eight tests. The contrast between the illuminance and humidity results is informative about the analysis as a whole. A design in which area-level disadvantage is associated with every quantity measured would invite the concern that the gradients reflect some feature of when, where, or by whom measurements were taken rather than genuine differences in environmental condition. That is not what we observe. Two of the four environmental exposures were clearly patterned, one was weakly patterned, and the control quantity was not patterned at all.

### 3.6. The noise pattern is not merely urban versus suburban

Because the pooled sample combined a dense urban core with rural municipalities, a natural concern is that the noise gradient simply reflects the contrast between loud, diverse cities and quieter, less diverse towns. The within-core analysis addresses this directly. Restricting the models to the 144 Providence-core sites, where the urban-suburban distinction does not apply but neighborhood composition still varies, the noise gradient persisted (Table 3, Supplementary Table S4). Noise remained significantly higher with the percentage of non-White residents (1.33 dBA, p = 0.014), the percentage of renters (1.45 dBA, p = 0.007), and the percentage of households without a vehicle (1.77 dBA, p < 0.001), and significantly lower with median income (p = 0.035) and the percentage of older residents (p = 0.032); the poverty association was retained in direction and of similar size but fell just short of significance (1.03 dBA, p = 0.057). Five of the eight markers remained significant within the urban core, and all retained their direction. The C-weighted levels behaved similarly within the core, remaining significantly higher with renter occupancy (1.46 dB, p = 0.005) and vehicle absence (1.53 dB, p = 0.003) and significantly lower with median income (−1.16 dB, p = 0.025), with racial composition and poverty retained in direction and magnitude but just short of significance (0.96 dB, p = 0.064; 0.98 dB, p = 0.059). The social gradient in the acoustic environment, under both weightings, was therefore present within the city itself, not only between the city and the rural municipalities.

**Table 3.** Noise associations across robustness specifications: primary (pooled, tract-level mixed model), site-mean ordinary least squares, block-group sensitivity (mixed model), and within- Providence-core (mixed model). Coefficients are standardized (per SD), in dBA.

| Neighborhood marker | Primary $\beta$ (p) | Site-mean OLS $\beta$ (p) | Block-group $\beta$ (p) | Within-core $\beta$ (p) |
| --- | --- | --- | --- | --- |

| **Neighborhood marker** | **Primary $\beta$ (p)** | **Site-mean OLS $\beta$ (p)** | **Block-group $\beta$ (p)** | **Within-core $\beta$ (p)** |
| --- | --- | --- | --- | --- |
| % non-White | 1.409 (0.017) | 1.425 (0.016) | 0.580 (0.310) | 1.326 (0.014) |
| % below 200% poverty | 1.256 (0.028) | 1.272 (0.026) | 0.695 (0.180) | 1.032 (0.057) |
| % less than high school | 0.497 (0.339) | 0.503 (0.334) | -0.483 (0.337) | 0.496 (0.361) |
| % renter-occupied | 1.772 (0.004) | 1.793 (0.004) | 2.201 (<0.001) | 1.454 (0.007) |
| % no vehicle | 1.746 (0.001) | 1.757 (0.001) | 1.717 (<0.001) | 1.774 (<0.001) |
| % under 18 | -0.810 (0.099) | -0.810 (0.100) | -0.668 (0.183) | -0.887 (0.102) |
| % over 65 | -1.540 (0.006) | -1.548 (0.006) | -1.811 (<0.001) | -1.161 (0.032) |
| Median household income | -1.342 (0.013) | -1.355 (0.013) | -0.765 (0.148) | -1.140 (0.035) |

### 3.7. Robustness

The findings were stable across specifications (Table 3). Collapsing each exposure to a single site-level mean and refitting with ordinary least squares reproduced the primary results almost exactly (Supplementary Table S2): the noise associations agreed in direction for all eight markers and in significance for six, with coefficients within a few hundredths of a decibel of the mixed-model estimates. Refitting at the block-group scale attenuated the associations, as expected given the larger margins of error of block-group estimates (Supplementary Table S3): three of the eight noise associations remained significant, with the percentage of renters, the percentage of households without a vehicle, and the percentage of older residents retaining strong and significant effects, while the weaker markers lost significance and, in the case of education, changed sign. This attenuation at finer geography is the expected signature of greater estimate uncertainty rather than evidence against the pattern, and the markers that survive are precisely those that are most reliably measured. The C-weighted results were similarly stable: the site-mean models reproduced the primary C-weighted estimates to within a few hundredths of a decibel, and, as noted above, the block-group models strengthened rather than attenuated the C-weighted associations with renter occupancy and vehicle absence. Across the primary, site-mean, within-core, and block-group analyses (summarized in Supplementary Table S5), the direction of the noise associations under both weightings was consistent for essentially every marker, and the strongest and most reliable associations, those with vehicle access, renter occupancy, race, and age, were significant in nearly every specification.

## 4. Discussion

Across 176 communities measured with the same protocol over roughly one year, environmental exposure was socially patterned, but not uniformly across the exposures we measured. More disadvantaged tracts, those with more non-White residents, more poverty, more renters, and fewer households with vehicles, were consistently louder under both A- and C-weighting, while wealthier and older neighborhoods were quieter. Night-time illuminance followed the same direction, with five of eight markers significant. Short-term PM2.5 leaned the same way but far more weakly, reaching significance for only two markers and with effects roughly an order of magnitude smaller, and relative humidity, the negative control, showed no coherent pattern. The acoustic gradient held within the Providence urban core alone and was stable across site-mean, block-group, and within-core analyses.

The most coherent reading of this contrast lies in the different physical natures of the two exposures. Noise is a property of place. It is dominated by road traffic and shaped by the configuration of roads, buildings, industry, and vegetation around a site, features that are fixed and that land-use regression models use to predict measured noise with considerable accuracy across cities [12,13,14,15]. Where disadvantaged communities are sited near busier roads, denser development, and less vegetation, as they often are, their higher noise is built into the surrounding fabric and persists. Short-term PM2.5 behaves differently. Much of the particulate burden in an urban area is regional and secondary, so concentrations tend toward spatial uniformity across a metropolitan area while changing from day to day with wind, precipitation, and atmospheric mixing [16,17]. A short measurement at a site therefore reflects the weather of that day across the wider area as much as the character of the location, which dilutes any spatial, and hence social, gradient that longer-term or modeled concentrations might reveal.

This interpretation also clarifies what our PM2.5 result does and does not mean. A large body of work has established that PM2.5 is socially patterned at the national scale, with people of color and lower-income populations exposed to systematically higher concentrations, and with these disparities widening over time [1,2,3]. Our finding is not in tension with that literature. The national studies rest on long-term, modeled, annual-average concentrations across many regions, whereas we measured short-term concentrations in a single small region over part of one year. The weak PM2.5 gradient we observed is most plausibly a consequence of that design, in which day-to-day meteorology dominates the measured signal, rather than evidence that particulate exposure is equitably distributed in Rhode Island. The directions of our PM2.5 associations, which align with the noise pattern, are if anything consistent with a faint underlying spatial gradient that our short-term sampling was not well suited to resolve. We would expect clearer social structure in settings we did not sample, such as areas with large local point sources, heavy curbside combustion, or meteorological regimes that trap pollution near its source, since it is the regional and secondary character of particulate mass that flattens within-region contrast [16,17]; our result speaks to a small region over a limited season, not to particulate inequality in general.

For the noise findings, our results extend a literature that has largely relied on modeled exposure. National and regional studies using modeled noise surfaces have repeatedly found higher noise in more disadvantaged and more heavily minority communities [18,19,20], and reviews report similar gradients internationally [21,22,23]. By measuring noise directly at each site, rather than assigning it from a model, we provide field confirmation that the gradient is present in observed sound levels and not only in modeled estimates, and we show it within a single urban core as well as across a wider region. The consistency of our gradient with the modeled-noise literature strengthens confidence in both.

The size of this gradient is not trivial in health terms. The exposure-response relationships underlying the WHO European noise guidelines are continuous and have no clear threshold below which transportation noise is harmless. Differences of a few decibels in long-term average exposure therefore shift the population burden of sleep disturbance and cardiovascular risk rather than leaving it unchanged [4,5,6,7,8]. A contrast of roughly 2 to 3 dBA across the interquartile range of neighborhood disadvantage, and of more than 6 dBA between the least and most racially diverse neighborhoods we sampled, therefore translates into a substantially greater chronic noise burden for communities that are already more disadvantaged. The importance of these differences lies not only in their magnitude but also in their unequal distribution: the communities with the highest measured noise were the same communities facing greater socioeconomic disadvantage, indicating that the burden of noise-related health risk is itself socially patterned.

The C-weighted results carry a distinct implication that the A-weighted analysis alone would not have surfaced. Because A-weighting discounts low-frequency energy, the conventional metric systematically understates the acoustic burden where that energy is abundant, and low-frequency sound is the component least attenuated by building facades and least amenable to barrier mitigation [24]. Our measurements show that the low-frequency component was not distributed independently of social composition: the C-weighted gradient ran in the same direction as the A-weighted gradient on every marker, was strongest for renter occupancy and vehicle absence, and at block-group scale exceeded the A-weighted associations for those two markers. The communities carrying the most low-frequency noise were, in this sample, the same communities with the least advantaged housing and transport circumstances. This matters practically because those are also the communities least able to remedy the exposure privately: low-frequency sound passes through the building envelope that would attenuate higher-frequency traffic noise, so the ordinary defenses of closed windows, insulation, and glazing offer less protection, and renters have limited ability to modify the envelope in any case. An assessment framework that describes community noise in A-weighted terms alone will therefore understate the burden precisely where it is greatest, and C-weighting, which is inexpensive to record on the same instrument at the same time, offers a direct remedy. The sources that generate low-frequency energy point in the same direction: heavy vehicle traffic on arterial roads, rail corridors including the Amtrak and commuter lines that pass through the urban core, industrial and mechanical plant, and, increasingly, distributed energy infrastructure such as wind turbines. Characterizing which of these dominate at a given site was beyond the scope of our protocol and is a natural next step.

The night-time illuminance result points the same way and reinforces the reading. Artificial light at night is associated with circadian disruption and shorter sleep, and that association has been found to be stronger in higher-poverty neighborhoods [25]. Finding measured illuminance elevated in the same tracts that carry higher measured noise indicates that two exposures capable of disturbing sleep, one acoustic and one visual, co-occur in the same places. Because both were measured at night with the same protocol, the co-occurrence is observed rather than inferred. Two cautions apply. Our illuminance measure is street-level rather than the satellite radiance used in most epidemiological studies of light at night, and it records total illuminance after dark rather than artificial light alone, so it is not directly comparable to that literature, and because illuminance is defined only for night sessions it rests on fewer observations than the acoustic results. We therefore treat it as a supporting finding rather than a co-primary one.

Taken together, these results support an environmental-justice reading of exposure in this setting, understood specifically as the unequal distribution of measured environmental burden across communities that differ in socioeconomic and racial composition. The evidence for that reading is stronger here than in studies that rely on a single modeled exposure, for three reasons. The exposures were measured rather than modeled, so the gradient is present in observed field data. The gradient appeared under two independent acoustic metrics and in a third, separately instrumented channel. And a quantity with no plausible link to neighborhood composition, relative humidity, showed no gradient, which argues against a methodological explanation for the pattern. At the same time, the specificity of the finding is as important as its existence: PM2.5 measured over short windows did not track composition, and any account of environmental burden in these communities that treated all exposures as interchangeable would have been wrong about which one was unequally distributed.

The clearest implication is for exposure assessment itself. The appeal of a composite environmental score, or of traffic proximity as a one-size surrogate, rests on the assumption that the burdens of a place travel together and can be summarized at once. In this setting they did not. A quiet site was not reliably a clean-air site, the social gradient was strong for noise and weak for short-term particulates, and the two exposures are governed by different processes, one spatial and persistent, the other temporal and weather-driven. Collapsing them into one index would blur a place-fixed inequality in noise with a weather-dominated particulate signal, and could direct attention away from the exposure that actually carried the disparity.

Two consequences follow. The first is misclassification: studies that use one exposure as a surrogate for the other, or that fold both into a single traffic or deprivation index, risk attributing to air pollution effects that belong to noise, or the reverse, because the two are organized along different axes and were here only weakly correlated. Studies of traffic-related health effects in socially patterned populations therefore have reason to measure noise and air separately rather than assume they travel together. The second concerns which disparities are visible at all. An inequality that is real but invisible to the dominant metric will not be acted on: if particulate concentrations are the only exposure routinely mapped, a noise disparity of the magnitude seen here would go unrecorded, and the communities bearing it would not be identified as overburdened. The same logic applies within a single exposure, because an assessment framework that reports A-weighted levels alone understates the acoustic burden precisely where low-frequency energy is most abundant.

Two design choices therefore carry more weight than they are usually given: the metric and the averaging time. Where resources allow only one exposure to be measured directly at fine spatial scale, our results suggest that noise is the more spatially informative and the more socially differentiated of the two in a compact, temperate region with no dominant local particulate source of the kind we sampled, while particulate inequality is better characterized with the long-term modeled estimates that the national literature relies on. Noise is also the more directly addressable, being tied to modifiable features of the built environment: road configuration and traffic management, sound-attenuating barriers and building standards, and tree canopy in the neighborhoods that currently bear the most noise. We did not record whether individual sites fell within areas subject to existing local noise measures such as municipal vehicle-noise ordinances, and linking measured levels to the presence and enforcement of such instruments would be a useful extension.

Several features of the design bound these conclusions. The study covers one small region over part of one year, so the specific coefficients invite replication rather than direct generalization, and the seasonal window samples a limited range of weather. Exposures were short spot measurements collected by many trained individuals rather than continuous monitor records. This tradeoff buys broad spatial coverage at the cost of long averaging at any one point, and it bears more heavily on PM2.5, whose signal is dominated by day-to-day weather, than on noise, whose short-sample measurements are known to track longer-term levels well [13]. As noted in the Methods, the particulate sensors support relative comparison rather than absolute concentration, so the PM2.5 analysis rests on relative structure across sites and times. Most fundamentally, our analysis is ecological. We linked exposures to the socioeconomic and racial composition of census tracts, not to the characteristics of the people measured, and our associations describe places rather than individuals. They are therefore subject to the ecological fallacy and to the modifiable areal unit problem, whereby estimates can shift with the size of the spatial unit chosen; the attenuation of our A-weighted results at the block-group scale is one illustration of the latter [31,32]. The margins of error of small-area ACS estimates compound this, and we have leaned on the more reliably measured composition markers and on tract-level rather than block-group geography for that reason [29,30]. These limitations temper the precision of our estimates but not the central, repeatedly reproduced contrast: across these communities, the social gradient in environmental exposure was carried by measured noise far more than by short-term measured particulates.

Environmental burden is often conceived as a single condition shared across multiple exposures. Our findings point to a more differentiated reality. In Rhode Island, directly measured noise under both A- and C-weighting, and measured night-time light, showed clear social gradients, whereas short-term measured PM2.5 did not, indicating that environmental burdens cannot always be inferred from one another. Characterizing environmental burden will therefore require measuring its constituent exposures individually, because what appears unequally distributed depends not only on where people live but also on what is measured.

## Declarations

### Ethics approval

This study analyzed environmental measurements collected in public outdoor locations together with publicly available aggregated census estimates. It did not involve human participants, and no individual-level or identifiable personal data were used; institutional review board approval was therefore not required.

### Data availability

The site-level environmental measurements and the derived analysis frame are available from the corresponding author on reasonable request. The American Community Survey estimates and the geographic reference files used for tract and block-group assignment are publicly available from the United States Census Bureau.

### Code availability

The analysis code used to generate the results reported here is available from the corresponding author on reasonable request.

### Author contributions

Erica D. Walker: Conceptualization, Data curation, Funding acquisition, Investigation, Methodology, Project administration, Resources, Software, Supervision, Validation, Writing - review & editing. Sai Venkat Mandalapu: Conceptualization, Data curation, Investigation, Methodology, Validation, Writing - original draft, Writing - review & editing. Sage Lefebvre: Conceptualization, Data curation, Formal analysis, Investigation, Methodology, Software, Validation, Visualization, Writing - review & editing.

## Acknowledgements

We thank the students of PHP1720 and PHP1721 at the Brown University School of Public Health for their work collecting the field measurements used in this study.

## Supplementary information

Supplementary Tables S1 to S8 are provided in a separate file.

## References

1. Tessum, C.W., Paolella, D.A., Chambliss, S.E., Apte, J.S., Hill, J.D., Marshall, J.D. PM2.5 polluters disproportionately and systemically affect people of color in the United States. Sci. Adv. 7, eabf4491 (2021).

2. Jbaily, A., Zhou, X., Liu, J., Lee, T.-H., Kamareddine, L., Verguet, S., Dominici, F. Air pollution exposure disparities across US population and income groups. Nature 601, 228– 233 (2022).

3. Kerr, G.H., van Donkelaar, A., Martin, R.V., Brauer, M., Bukart, K., Wozniak, S., Goldberg, D.L., Anenberg, S.C. Increasing racial and ethnic disparities in ambient air pollution-attributable morbidity and mortality in the United States. Environ. Health Perspect. 132, 037002 (2024).

4. World Health Organization Regional Office for Europe. Environmental Noise Guidelines for the European Region. WHO Regional Office for Europe, Copenhagen (2018).

5. van Kempen, E., Casas, M., Pershagen, G., Foraster, M. WHO environmental noise guidelines for the European region: a systematic review on environmental noise and cardiovascular and metabolic effects. Int. J. Environ. Res. Public Health 15, 379 (2018).

6. Münzel, T., Sørensen, M., Daiber, A. Transportation noise pollution and cardiovascular disease. Nat. Rev. Cardiol. 18, 619–636 (2021).

7. Basner, M., McGuire, S. WHO environmental noise guidelines for the European region: a systematic review on environmental noise and effects on sleep. Int. J. Environ. Res. Public Health 15, 519 (2018).

8. Smith, M.G., Cordoza, M., Basner, M. Environmental noise and effects on sleep: an update to the WHO systematic review and meta-analysis. Environ. Health Perspect. 130, 076001 (2022).

9. Allen, R.W., Davies, H., Cohen, M.A., Mallach, G., Kaufman, J.D., Adar, S.D. The spatial relationship between traffic-generated air pollution and noise in 2 US cities. Environ. Res. 109, 334–342 (2009).

10. Tétreault, L.-F., Perron, S., Smargiassi, A. Cardiovascular health, traffic-related air pollution and noise: are associations mutually confounded? A systematic review. Int. J. Public Health 58, 649–666 (2013).

11. Wild, C.P. Complementing the genome with an “exposome”: the outstanding challenge of environmental exposure measurement in molecular epidemiology. Cancer Epidemiol. Biomarkers Prev. 14, 1847–1850 (2005).

12. Aguilera, I., Foraster, M., Basagaña, X., Corradi, E., Deltell, A., Morelli, X., Phuleria, H.C., Ragettli, M.S., Rivera, M., Thomasson, A., Slama, R., Künzli, N. Application of land use regression modelling to assess the spatial distribution of road traffic noise in three European cities. J. Expo. Sci. Environ. Epidemiol. 25, 97–105 (2015).

13. Ragettli, M.S., Goudreau, S., Plante, C., Fournier, M., Hatzopoulou, M., Perron, S., Smargiassi, A. Statistical modeling of the spatial variability of environmental noise levels in Montreal, Canada, using noise measurements and land use characteristics. J. Expo. Sci. Environ. Epidemiol. 26, 597–605 (2016).

14. Staab, J., Schady, A., Weigand, M., Lakes, T., Taubenböck, H. Predicting traffic noise using land-use regression: a scalable approach. J. Expo. Sci. Environ. Epidemiol. 32, 232–243 (2022).

15. Yin, X., Fallah-Shorshani, M., McConnell, R., Fruin, S., Franklin, M. Predicting fine spatial scale traffic noise using mobile measurements and machine learning. Environ. Sci. Technol. 54, 12860–12869 (2020).

16. Pinto, J.P., Lefohn, A.S., Shadwick, D.S. Spatial variability of PM2.5 in urban areas in the United States. J. Air Waste Manage. Assoc. 54, 440–449 (2004).

17. Tai, A.P.K., Mickley, L.J., Jacob, D.J. Correlations between fine particulate matter (PM2.5) and meteorological variables in the United States: implications for the sensitivity of PM2.5 to climate change. Atmos. Environ. 44, 3976–3984 (2010).

18. Casey, J.A., Morello-Frosch, R., Mennitt, D.J., Fristrup, K., Ogburn, E.L., James, P. Race/ethnicity, socioeconomic status, residential segregation, and spatial variation in noise exposure in the contiguous United States. Environ. Health Perspect. 125, 077017 (2017).

19. Collins, T.W., Grineski, S.E. Race, historical redlining, and contemporary transportation noise disparities in the United States. J. Expo. Sci. Environ. Epidemiol. 35, 50–61 (2025).

20. Shkembi, A., Patel, K., Smith, L.M., Meier, H.C.S., Neitzel, R.L. Racial and ethnic inequities to noise pollution from transportation- and work-related sources in the United States. J. Expo. Sci. Environ. Epidemiol. 36, 211–220 (2026).

21. Dreger, S., Schüle, S.A., Hilz, L.K., Bolte, G. Social inequalities in environmental noise exposure: a review of evidence in the WHO European Region. Int. J. Environ. Res. Public Health 16, 1011 (2019).

22. Trudeau, C., King, N., Guastavino, C. Investigating sonic injustice: a review of published research. Soc. Sci. Med. 326, 115927 (2023).

23. Klompmaker, J.O., Hoek, G., Bloemsma, L.D., Gehring, U., Strak, M., Wijga, A.H., Janssen, N.A.H. Environmental noise is positively associated with socioeconomically less privileged neighborhoods in the Netherlands. Environ. Res. 248, 118219 (2024).

24. Leventhall, H.G. Low frequency noise and annoyance. Noise Health 6 (23), 59–72 (2004).

25. Xiao, Q., Gee, G., Jones, R.R., Jia, P., James, P., Hale, L. Cross-sectional association between outdoor artificial light at night and sleep duration in middle-to-older aged adults: The NIH-AARP Diet and Health Study. Environ. Res. 180, 108823 (2020).

26. Rhode Island Department of Health. Rural Definition. Rhode Island Department of Health, Providence, RI. https://health.ri.gov/sites/g/files/xkgbur1006/files/publications/definitions/2022Rural-definition.pdf (2022).

27. Centers for Disease Control and Prevention. Asthma Surveillance in the United States, 2001-2021. National Center for Environmental Health, Centers for Disease Control and Prevention, Atlanta, GA (2023).

28. Rhode Island Department of Environmental Management. Environmental Justice. Rhode Island Department of Environmental Management, Providence, RI. https://dem.ri.gov/environmental-protection-bureau/initiatives/environmental-justice (2024).

29. Spielman, S.E., Folch, D., Nagle, N. Patterns and causes of uncertainty in the American Community Survey. Appl. Geogr. 46, 147–157 (2014).

30. Donegan, C., Chun, Y., Griffith, D.A. Modeling community health with areal data: Bayesian inference with survey standard errors and spatial structure. Int. J. Environ. Res. Public Health 18, 6856 (2021).

31. Openshaw, S. Ecological fallacies and the analysis of areal census data. Environ. Plan. A 16, 17–31 (1984).

32. Parenteau, M.-P., Sawada, M.C. The modifiable areal unit problem (MAUP) in the relationship between exposure to NO2 and respiratory health. Int. J. Health Geogr. 10, 58 (2011).

