## Supplementary Materials for "Divergent social patterning of directly measured environmental exposures across Rhode Island communities"

This document provides the full set of coefficients underlying the analyses summarized in the main text. It reports the complete results of the primary pooled tract-level models for both exposures (Supplementary Table S1), the site-mean ordinary least squares models (Supplementary Table S2), the block-group sensitivity models (Supplementary Table S3), the within-Providence-core models (Supplementary Table S4), and a cross-specification consistency summary for noise (Supplementary Table S5). All analyses use the same 176 sites described in the main text. Throughout, the acoustic outcome is the A-weighted equivalent continuous sound level (LAeq, in dBA) and the air-quality outcome is log-transformed fine particulate matter (log PM2.5). Each neighborhood composition marker was standardized to zero mean and unit variance and entered as the sole predictor in its own model, so coefficients are interpreted per standard deviation of the marker. Unless stated otherwise, models include a random intercept for site, a fixed effect for session type (weekday daytime as reference), and a fixed effect for campaign.

**S1. Primary specification: pooled, tract-level, campaign- and session-adjusted mixed models**

Supplementary Table S1 reports the standardized coefficient, standard error, and p-value for each of the eight neighborhood composition markers, for both noise and PM2.5, in the primary specification. These are the full results summarized in Table 2 and Figure 1 of the main text. For noise, six of the eight markers reach conventional significance, and all eight are oriented so that greater disadvantage corresponds to higher noise. For PM2.5, the associations follow the same directions but are roughly an order of magnitude smaller in standardized terms, and only the percentage below 200% of poverty and the percentage of households without a vehicle reach significance.

**Supplementary Table S1.** Primary pooled tract-level mixed models. Standardized coefficients (per SD), standard errors, and p-values for noise (LAeq) and log PM2.5.

| Neighborhood marker | Noise β | Noise SE | Noise p | PM2.5 β | PM2.5 SE | PM2.5 p |
| --- | --- | --- | --- | --- | --- | --- |
| % non-White | 1.409 | 0.586 | 0.017 | 0.051 | 0.027 | 0.057 |
| % below 200% poverty | 1.256 | 0.567 | 0.028 | 0.060 | 0.025 | 0.020 |
| % less than high school | 0.497 | 0.518 | 0.339 | 0.009 | 0.023 | 0.716 |
| % renter-occupied | 1.772 | 0.613 | 0.004 | 0.051 | 0.028 | 0.071 |
| % no vehicle | 1.746 | 0.527 | 0.001 | 0.054 | 0.024 | 0.025 |
| % under 18 | -0.810 | 0.488 | 0.099 | -0.010 | 0.022 | 0.666 |
| % over 65 | -1.540 | 0.556 | 0.006 | -0.034 | 0.026 | 0.196 |
| Median household income | -1.342 | 0.536 | 0.013 | -0.044 | 0.024 | 0.069 |

**S2. Site-mean ordinary least squares models**

To confirm that the findings are not produced by the repeated-measures structure of the data, each exposure was collapsed to a single mean per site and the eight models refitted with ordinary least squares, using the same standardized composition markers and adjusting for campaign. Supplementary Table S2 reports the results. The site-mean coefficients reproduce the primary mixed-model coefficients almost exactly for noise, agreeing in direction for all eight markers and in significance for six, which confirms that the noise gradient is a property of the site-level exposure and not an artifact of the within-site repeated measurements. The PM2.5 associations are likewise close to the primary estimates.

**Supplementary Table S2.** Site-mean ordinary least squares models. Standardized coefficients (per SD) and p-values for noise and log PM2.5.

| Neighborhood marker | Noise β | Noise p | PM2.5 β | PM2.5 p |
| --- | --- | --- | --- | --- |
| % non-White | 1.425 | 0.016 | 0.069 | 0.015 |
| % below 200% poverty | 1.272 | 0.026 | 0.075 | 0.006 |
| % less than high school | 0.503 | 0.334 | 0.017 | 0.504 |
| % renter-occupied | 1.793 | 0.004 | 0.059 | 0.049 |
| % no vehicle | 1.757 | 0.001 | 0.050 | 0.056 |
| % under 18 | -0.810 | 0.100 | 0.004 | 0.859 |
| % over 65 | -1.548 | 0.006 | -0.059 | 0.030 |
| Median household income | -1.355 | 0.013 | -0.040 | 0.128 |

**S3. Block-group sensitivity models**

To test the dependence of the findings on the spatial scale of the neighborhood unit, the primary models were refitted using block-group rather than census-tract composition. Supplementary Table S3 reports the results. The noise associations attenuate at the block-group scale, which is the expected consequence of the larger margins of error of block-group ACS estimates relative to tract estimates. Three of the eight noise associations remain significant: the percentage of renters, the percentage of households without a vehicle, and the percentage of older residents, which are among the more reliably measured markers. The percentage with less than a high-school education changes sign at this scale but remains non-significant. This pattern of attenuation, concentrated in the less reliably measured markers, is consistent with the modifiable areal unit problem and with measurement uncertainty rather than with an absence of the underlying gradient.

**Supplementary Table S3.** Block-group sensitivity mixed models. Standardized coefficients (per SD), standard errors, and p-values for noise and log PM2.5.

| **Neighborhood marker** | **Noise β** | **Noise SE** | **Noise p** | **PM2.5 β** | **PM2.5 SE** | **PM2.5 p** |
| --- | --- | --- | --- | --- | --- | --- |
| % non-White | 0.580 | 0.570 | 0.310 | 0.021 | 0.026 | 0.408 |
| % below 200% poverty | 0.695 | 0.516 | 0.180 | 0.003 | 0.023 | 0.912 |
| % less than high school | -0.483 | 0.502 | 0.337 | -0.019 | 0.023 | 0.395 |
| % renter-occupied | 2.201 | 0.558 | <0.001 | 0.028 | 0.026 | 0.289 |
| % no vehicle | 1.717 | 0.499 | <0.001 | -0.008 | 0.024 | 0.746 |
| % under 18 | -0.668 | 0.500 | 0.183 | 0.000 | 0.023 | 0.999 |
| % over 65 | -1.811 | 0.507 | <0.001 | -0.047 | 0.024 | 0.050 |
| Median household income | -0.765 | 0.527 | 0.148 | 0.019 | 0.024 | 0.423 |

**S4. Within-Providence-core models**

To test whether the noise gradient reflects only the contrast between the dense urban core and the rural municipalities, the primary models were refitted using the 144 sites in the Providence urban core alone. Within the core, the urban-suburban distinction does not apply, but neighborhood composition still varies substantially. Supplementary Table S4 reports the results. The noise gradient persists: five of the eight markers remain significant, and all eight retain the direction seen in the pooled analysis, with greater disadvantage corresponding to higher noise. The poverty association is retained in direction and magnitude but falls just short of conventional significance. This indicates that the social gradient in noise is present within the city itself and is not an artifact of pooling the two campaigns.

Supplementary Table S4. Within-Providence-core mixed models (144 sites). Standardized coefficients (per SD), standard errors, and p-values for noise and log PM2.5.

| **Neighborhood marker** | **Noise β** | **Noise SE** | **Noise p** | **PM2.5 β** | **PM2.5 SE** | **PM2.5 p** |
| --- | --- | --- | --- | --- | --- | --- |
| % non-White | 1.326 | 0.532 | 0.014 | 0.046 | 0.025 | 0.073 |
| % below 200% poverty | 1.032 | 0.537 | 0.057 | 0.053 | 0.025 | 0.034 |
| % less than high school | 0.496 | 0.542 | 0.361 | 0.011 | 0.025 | 0.678 |
| % renter-occupied | 1.454 | 0.530 | 0.007 | 0.043 | 0.025 | 0.090 |
| % no vehicle | 1.774 | 0.523 | <0.001 | 0.057 | 0.025 | 0.023 |
| % under 18 | -0.887 | 0.538 | 0.102 | -0.011 | 0.026 | 0.671 |
| % over 65 | -1.161 | 0.534 | 0.032 | -0.025 | 0.026 | 0.337 |
| Median household income | -1.140 | 0.535 | 0.035 | -0.040 | 0.025 | 0.108 |

**S5. Cross-specification consistency for noise**

Supplementary Table S5 places the noise coefficient and p-value for each marker side by side across the four specifications and records whether the direction of the association agrees with the primary specification. The direction of the noise association agrees with the primary model in every specification for every marker, with the single exception of the percentage with less than a high-school education at the block-group scale, which is the least reliably measured marker and is non-significant throughout. The strongest and most reliably measured associations, those with vehicle access, renter occupancy, race, and the percentage of older residents, are significant in nearly every specification.

**Supplementary Table S5.** Cross-specification consistency for noise. Standardized noise coefficients (per SD) and p-values across the primary pooled tract model, site-mean OLS, block-group, and within-core specifications, with direction-agreement flags relative to the primary specification.

| **Marker** | **Primary β (p)** | **OLS β (p)** | **Block-group β (p)** | **Within-core β (p)** | **Dir. agree** |
| --- | --- | --- | --- | --- | --- |
| % non-White | 1.409 (0.017) | 1.425 (0.016) | 0.580 (0.310) | 1.326 (0.014) | yes / yes / yes |
| % below 200% poverty | 1.256 (0.028) | 1.272 (0.026) | 0.695 (0.180) | 1.032 (0.057) | yes / yes / yes |
| % less than high school | 0.497 (0.339) | 0.503 (0.334) | -0.483 (0.337) | 0.496 (0.361) | yes / no / yes |
| % renter-occupied | 1.772 (0.004) | 1.793 (0.004) | 2.201 (<0.001) | 1.454 (0.007) | yes / yes / yes |
| % no vehicle | 1.746 (0.001) | 1.757 (0.001) | 1.717 (<0.001) | 1.774 (<0.001) | yes / yes / yes |
| % under 18 | -0.810 (0.099) | -0.810 (0.100) | -0.668 (0.183) | -0.887 (0.102) | yes / yes / yes |
| % over 65 | -1.540 (0.006) | -1.548 (0.006) | -1.811 (<0.001) | -1.161 (0.032) | yes / yes / yes |
| Median household income | -1.342 (0.013) | -1.355 (0.013) | -0.765 (0.148) | -1.140 (0.035) | yes / yes / yes |

Direction-agreement columns refer to site-mean OLS / block-group / within-core, each relative to the primary specification. The single non-agreement is the percentage with less than a high-school education at the block-group scale, the least reliably measured marker, which is non-significant throughout.

**Note on the air-quality (PM2.5) associations across specifications**

For completeness, the PM2.5 associations can be compared across the primary (S1), site-mean (S2), block-group (S3), and within-core (S4) specifications. Across these, the PM2.5 associations are consistently small in standardized terms and reach significance for at most a few markers in any given specification (most often the percentage below 200% of poverty and the percentage of households without a vehicle). Their directions broadly track the noise associations, which is consistent with both exposures sharing traffic as a partial source, but their magnitude relative to noise, and their inconsistency across specifications, support the main-text conclusion that the social patterning of exposure in this setting is carried mainly by the acoustic environment rather than by short-term measured particulate concentrations.

**S6. C-weighted noise (LCeq) associations across specifications**

Standardized associations between neighborhood composition and C-weighted noise, expressed as beta (p) in dB per standard deviation of the composition marker. Specifications match those used for A-weighted noise in Tables 2 and 3.

| **Neighborhood marker** | **Primary** | **Site-mean OLS** | **Block-group** | **Within-core** |
| --- | --- | --- | --- | --- |
| % non-White | 1.069 (0.053) | 1.083 (0.051) | 0.558 (0.295) | 0.959 (0.064) |
| % below 200% poverty | 1.060 (0.047) | 1.074 (0.045) | 0.840 (0.082) | 0.978 (0.059) |
| % less than high school | 0.375 (0.439) | 0.381 (0.433) | -0.179 (0.704) | 0.440 (0.397) |
| % renter-occupied | 1.662 (0.004) | 1.681 (0.004) | 2.354 (<0.001) | 1.458 (0.005) |
| % no vehicle | 1.513 (0.003) | 1.523 (0.002) | 1.938 (<0.001) | 1.531 (0.003) |
| % under 18 | -0.467 (0.309) | -0.466 (0.310) | -0.631 (0.178) | -0.522 (0.315) |
| % over 65 | -0.750 (0.156) | -0.757 (0.153) | -1.216 (0.012) | -0.615 (0.236) |
| Median household income | -1.156 (0.022) | -1.168 (0.021) | -0.858 (0.086) | -1.156 (0.025) |

**S7. Z-weighted noise (LZeq) associations across specifications**

Standardized associations between neighborhood composition and unweighted (Z-weighted) noise, expressed as beta (p) in dB per standard deviation. Reported for completeness; the main text relies on the A- and C-weighted metrics.

| **Neighborhood marker** | **Primary** | **Site-mean OLS** | **Block-group** | **Within-core** |
| --- | --- | --- | --- | --- |
| % non-White | 0.910 (0.086) | 0.926 (0.081) | 0.436 (0.394) | 0.801 (0.103) |
| % below 200% poverty | 0.819 (0.111) | 0.835 (0.104) | 0.645 (0.164) | 0.840 (0.087) |
| % less than high school | 0.302 (0.515) | 0.309 (0.507) | -0.321 (0.476) | 0.388 (0.431) |
| % renter-occupied | 1.635 (0.003) | 1.655 (0.003) | 2.392 (<0.001) | 1.518 (0.002) |
| % no vehicle | 1.621 (<0.001) | 1.631 (<0.001) | 1.992 (<0.001) | 1.634 (<0.001) |
| % under 18 | -0.520 (0.236) | -0.520 (0.237) | -0.743 (0.097) | -0.583 (0.236) |
| % over 65 | -0.450 (0.375) | -0.458 (0.368) | -0.868 (0.063) | -0.474 (0.336) |
| Median household income | -0.993 (0.041) | -1.006 (0.038) | -0.615 (0.199) | -1.144 (0.019) |

**S8. Night-time illuminance and relative humidity associations**

Standardized associations between neighborhood composition and night-time outdoor illuminance (footcandles per standard deviation) and relative humidity (percentage points per standard deviation), for the primary pooled specification and the within-Providence-core specification. Humidity is included as a negative control.

| **Neighborhood marker** | **Illuminance, primary** | **Illuminance, within-core** | **Humidity, primary** | **Humidity, within-core** |
| --- | --- | --- | --- | --- |
| % non-White | 0.193 (0.006) | 0.183 (0.005) | 1.076 (0.094) | 0.970 (0.060) |
| % below 200% poverty | 0.240 (<0.001) | 0.208 (0.002) | 0.662 (0.281) | 1.112 (0.029) |
| % less than high school | 0.155 (0.018) | 0.159 (0.022) | 0.500 (0.372) | 0.725 (0.160) |
| % renter-occupied | 0.126 (0.111) | 0.105 (0.134) | 0.199 (0.766) | 0.568 (0.266) |
| % no vehicle | 0.073 (0.325) | 0.072 (0.335) | -0.077 (0.894) | 0.089 (0.860) |
| % under 18 | 0.013 (0.844) | 0.023 (0.758) | 1.167 (0.027) | 1.349 (0.008) |
| % over 65 | -0.148 (0.027) | -0.108 (0.102) | 0.156 (0.803) | -0.375 (0.480) |
| Median household income | -0.144 (0.038) | -0.126 (0.074) | -0.424 (0.463) | -0.819 (0.105) |
